# Spatiotemporal transmission of SARS-CoV-2 lineages during 2020-2021 in Pernambuco - Brazil

**DOI:** 10.1101/2023.01.25.23284831

**Authors:** Lais Ceschini Machado, Filipe Zimmer Dezordi, Gustavo Barbosa de Lima, Raul Emídio de Lima, Lilian Caroliny Amorim Silva, Leandro de Mattos Pereira, Alexandre Freitas da Silva, Antonio Marinho da Silva Neto, André Luiz Sá de Oliveira, Anderson da Costa Armstrong, Rômulo Pessoa-e-Silva, Rodrigo Moraes Loyo, Barbara de Oliveira Silva, Anderson Rodrigues de Almeida, Maira Galdino da Rocha Pitta, Francisco de Assis da Silva Santos, Marilda Mendonça Siqueira, Paola Cristina Resende, Edson Delatorre, Felipe Gomes Naveca, Fabio Miyajima, Tiago Gräf, Rodrigo Feliciano do Carmo, Michelly Cristiny Pereira, Tulio de Lima Campos, Matheus Filgueira Bezerra, Marcelo Henrique Santos Paiva, Gabriel da Luz Wallau, Fiocruz COVID-19 Genomic Network

**Affiliations:** Departamento de Entomologia, Instituto Aggeu Magalhães (IAM)-Fundação Oswaldo Cruz-FIOCRUZ, Recife 50670-420, Brazil; Núcleo de Plataformas Tecnológicas (NPT), Instituto Aggeu Magalhães (IAM), FIOCRUZ-Pernambuco, Recife, Pernambuco, Brazil; Núcleo de Bioinformática (NBI), Instituto Aggeu Magalhães (IAM), FIOCRUZ-Pernambuco, Recife, Pernambuco, Brazil; Núcleo de Estatística e Geoprocessamento, Instituto Aggeu Magalhães (IAM)- Fundação Oswaldo Cruz Pernambuco- FIOCRUZ-PE, Recife 50670-420, Brazil; Colegiado de Ciências Farmacêuticas, Universidade Federal do Vale do São Francisco, Petrolina, Brasil; Colegiado de Medicina, Universidade Federal do Vale do São Francisco, Petrolina, Brasil; Suely-Galdino Therapeutic Innovation Research Center (NUPIT-SG). Federal University of Pernambuco (UFPE), Av. Professor Moraes Rêgo, s/n, Pernambuco, Brazil. CEP: 50670-901; Departamento de Parasitologia, Instituto Aggeu Magalhães (IAM), FIOCRUZ-Pernambuco, Recife, Pernambuco, Brazil; Núcleo de Ciências da Vida, Universidade Federal de Pernambuco (UFPE), Centro Acadêmico do Agreste-Rodovia BR-104, km 59-Nova Caruaru, Caruaru 55002-970, Brazil; Laboratory of Respiratory Viruses and Measles (LVRS), Instituto Oswaldo Cruz, FIOCRUZ-Rio de Janeiro, Rio de Janeiro, Brazil; Departamento de Biologia. Centro de Ciências Exatas, Naturais e da Saúde, Universidade Federal do Espírito Santo, Alegre, Espírito Santo, Brazil; Laboratório de Ecologia de Doenças Transmissíveis na Amazônia (EDTA), Instituto Leônidas e Maria Deane, FIOCRUZ-Amazonas, Manaus, Amazonas, Brazil; Analytical Competence Molecular Epidemiology Laboratory (ACME), FIOCRUZ-Ceará, Fortaleza, Ceará, Brazil; Laboratório de Virologia Molecular, Instituto Carlos Chagas, Fundação Oswaldo Cruz, Curitiba, Paraná, Brasil; Departamento de Microbiologia, Instituto Aggeu Magalhães (IAM), FIOCRUZ-Pernambuco, Recife, Pernambuco, Brazil; Wellcome Sanger Institute · Genomic Survaillance Unit, Wellcome Trust Genome Campus, Hinxton, Saffron Walden CB10 1RQ, Reino Unido; Department of Arbovirology, Bernhard Nocht Institute for Tropical Medicine, WHO Collaborating Center for Arbovirus and Hemorrhagic Fever Reference and Research. National Reference Center for Tropical Infectious Diseases. Bernhard-Nocht-Straße 74, 20359 Hamburg - Germany

**Keywords:** genomic surveillance, COVID-19, lineage replacement, state level

## Abstract

In recent years, the SARS-CoV-2 viruses emerged and spread around the world, leaving a large death toll and long-lasting impact on survivors. As of January 2023, Brazil is still among the countries with the highest number of registered deaths. This continental-size and pluralistic country experienced a heterogenous implementation of non-pharmacological and pharmacological interventions which, associated with large socioeconomic differences between the country regions, has led to distinct virus spread dynamics across the country. Here we investigate the spatiotemporal dispersion of emerging SARS-CoV-2 lineages and its dynamics in distinct epidemiological scenarios in the first two years of the pandemics in the Pernambuco state (Northeast Brazil). We generated a total of 1389 new SARS-CoV-2 genomes from June 2020 to August 2021 covering all major regions of the state. This sampling captured the arrival, communitary transmission and the circulation of the B1.1, B.1.1.28 and B.1.1.33 lineages in the first eight months of the pandemics, the emergence of the former variant of interest P.2 and the emergence and fast replacement of all previous variants by the more transmissible variant of concern P.1 (Gamma) lineage. Based on the incidence and lineage spread pattern we observed that there was an East-to-West to inner state pattern of transmission which is in agreement with the transmission of more populous metropolitan areas to medium and small size country-side cities in the state. Such transmission patterns may be partially explained by the main routes of traffic across municipalities in the state. Nevertheless, inter-state traffic was also another important source of lineage introduction and spread into the state. Our results highlight that the fine grained intrastate analysis of lineages and incidence spread can provide actionable insights for planning future non-pharmacological intervention for air-borne transmissible human pathogens.

## Introduction

Sustained SARS-CoV-2 human transmission was first characterized as an unknown/unusual pneumonia in the province of Wuhan in China in December 2019[1]. Since then, this virus has spread worldwide causing the largest pandemic recorded in the last 100 years. To date (November 2022) up to 628 million diagnosed cases are known and more than 6 million deaths were registered worldwide[2]. Due to recent technological and scientific advances on genome sequencing and bioinformatic analysis, it has been possible to closely monitor the SARS-CoV-2 evolution and track in almost real-time the emergence and replacement dynamics of Variants of Concern (VOCs)[3]. VOCs carry a combination of Spike amino acid changes and indels that enhance receptor binding to human ACE2 receptors and/or provide immune escape capacity from specific neutralizing antibodies[4–6] besides many other mutations along the genome which are poorly studied so far[7].

The world experienced the emergence of the first generation of VOCs by the end of 2020, such as Alpha (B.1.1.7), Beta (B.1.351) and Gamma (P.1), which had variable spread among countries, but all caused massive surges in cases mostly in their countries of origin[8–10]. It was then followed by second generation of VOCs, such as Delta and Omicron, which were more transmissible and carried a large array of immune escape mutations [9–11]. Both lineages are substantially more transmissible than previous circulating lineages and rapidly replaced them[11–14]. However, the replacement dynamics of previous lineages by the Delta variant had a rather heterogeneous impact on the increase of new cases in distinct regions of the world. While a direct association between Delta increasing frequency and sustained increment in diagnosed cases was well established in European countries, in Brazil, Delta replacement occurred during a sustained decrease in cases[15,16]. On the other hand, Omicron has shown a unique immune escape profile and has spread globally replacing Delta and increasing the infection rate in all countries[17]. In Brazil, a combination of several factors, such as: more transmissible VOCs (most notably Gamma and Omicron); limited application and engagement to non-pharmacological control measures; and a systematically weakened public health system lamentably resulted in over 34 million cases and at least 680 thousand deaths up to November 2022[2].

Genomic surveillance in Brazil was limited during the beginning of the pandemic, but specific initiatives successfully detected and characterized the international introduction of lineages and community transmission of distinct lineages as well as the introduction and spread of VOCs[18–20]. With the establishment of research networks, SARS-CoV-2 surveillance has improved substantially in the country, providing country-wide views of lineage introduction, spread, establishment and transmission patterns between states[21–24]. But, although interstate SARS-CoV-2 dynamics were more thoroughly investigated, up to October 2022 there were limited intrastate SARS-CoV-2 studies. Only Amazonas, Santa Catarina, Tocantins, Paraná, São Paulo and Rio de Janeiro intrastate dynamics have been investigated[21,24–28] as well as some reports from Rio Grande do Sul and Minas Gerais states focussing on specific lineage replacements[18,29,30]. Our group published the first genomes from Pernambuco state showing multiple introductions and ongoing community transmission at the beginning of the pandemic in Brazil[31]. Interstate transmission has been investigated for specific lineages such as P.1 Gamma and P.2 lineages in Pernambuco[32]. Although these studies provided crucial information to understand the SARS-CoV-2 dynamics in Brazil, states or region-specific SARS-CoV-2 lineage dynamics are expected due to the large differences between inter and intra states population sizes, concentration and human movement dynamics in Brazil[21]. In addition, non-pharmacological strategies to control SARS-CoV-2 were implemented with highly distinct adoption patterns across the country[33]. Therefore, understanding the dynamics of SARS-CoV-2 within the individual states is of paramount importance to understand the main determinants of virus spread grounded on specific state characteristics at a fine-grained resolution.

We sequenced 1,389 SARS-CoV-2 genomes from Pernambuco state in this retrospective study. Analyzing the new dataset plus the available genomic dataset from the state, we investigate the COVID-19 pandemic in the state in a lineage transmission dynamics perspective from March 2020 to early August 2021. We describe the timing and spread dynamics of different infection waves in Pernambuco, considering geographical traffic data from federal and state highways as a proxy of human mobility within the state. Our results show that the rapid spread of both VOCs and non-VOCs lineages is driven by more populous urban centers located in the east and west sides of Pernambuco, towards the inner and less populated counties. As well as the principal intra-state highway traffic mirrors this spread pattern and was likely a key component of SARS-CoV-2 lineages dissemination in the state.

## Material and Methods

### Genomic sequencing

Nasopharyngeal samples were obtained from two partnerships with Laboratório Central de Saúde Pública de Pernambuco (LACEN-PE) and the Núcleo de Pesquisa em Inovação Terapêutica - Suely Galdino (NUPIT-SG). RT-qPCR tests were performed using Kit Biomol OneStep Covid-19 (IBMP) and Kit Molecular SARS-CoV-2 (Bio-manguinhos) according to the manufacturer. Samples with Ct < 25 were further processed for amplification and sequencing. During the study period we employed three different methodologies to generate cDNA and amplify the SARS-CoV-2 genome: short amplicon according to Quick, 2020 (version 1); long amplicon according to Resende, 2020[34]; and inserting three sets of primers in COVIDSeq protocol according to Naveca 2021[35]. Genome-wide amplicons were then processed for sequencing using Nextera XT (Illumina, San Diego, CA, USA) or COVIDSeq (Illumina, San Diego, CA, USA) library preparation protocols following manufacturer instructions. Sequencing was performed using the MiSeq employing MiSeq Reagent kit V3 - paired-end 150 cycles flow cell. The ethics committee was approved by CEP FIOCRUZ/IAM - CAAE 32333120.4.0000.5190.

### Genome Assembly and Annotation

A reference-guided genome assembly strategy was employed using ViralFlow[36]. Briefly low-quality reads were trimmed in sliding window mode of 4 bases with a mean phred score threshold of 20, PCR primers input and adaptors were removed, and a minimum of 5x coverage depth of bases with Phred score quality equals to 30 were used to call a base to the major consensus genome.

### Spike region genotyping analysis with Sanger sequencing

We performed a Spike region genotyping of 135 additional SARS-CoV-2 RT-qPCR positive samples between January and April 2021, using a mutation profile of the RBD region of Spike protein according to Bezerra 2021[37], updated with the primer set described at flanking 22607-23446 region, (1MS Fw 5’-TAACGCCACCAGATTTGCAT-3’, 2MS Rv 5’-ACACGCCAAGTAGGAGTAAGT-3’) (dx.doi.org/10.17504/protocols.io.ewov1nxqkgr2/v2).

### Temporal and Phylodynamic analysis

We recovered the full dataset of 3,201,025 SARS-CoV-2 genomes from the GISAID database (https://www.gisaid.org/) on September 2, 2021. Subsequently, we performed a random subsampling with Augur 6.3.0 (https://docs.nextstrain.org/projects/augur/en/stable/index.html) pipeline obtaining 7,228 genome sequences (minimum 93% coverage) containing representative genomes from all continents and with focal context including 1,587 genomes from Pernambuco state where 1,490 were sequenced by the Aggeu Magalhães Institute at Fiocruz. The 7,228 genomes were aligned against the Wuhan-Hu-1 reference genome (GenBank NC_045512.2, GISAID EPI_ISL_402125) using MAFFT v7.47152 then the UTR regions were masked, and the phylogenetic tree was reconstructed with the Neighbor Joining method implemented in Augur. An additional phylogenetic tree was reconstructed with IQ-TREE2[38] using maximum likelihood with the model selected with ModelFinder[39] and SH-aLRT support values calculated from 1000 replicates.

The temporal spreading pattern of the identified SARS-CoV-2 monophyletic clades was analyzed firstly using the maximum likelihood approach and microreact (https://microreact.org/project/k9UFJmkrZHSx9RmFbDwZah-sars-cov-2-final-update-pernambuco). To understand how many clades were in circulation in the Pernambuco state the initial phylogeny of 7,228 genomes was manually inspected, the monophyletic clades with 10 or more genomes from Pernambuco showing clade support score (aLRT) higher than 80 were recovered. The resulting dataset with 258 genomes was submitted to a Bayesian phylogenetic analysis with the Markov Chain Monte Carlo (MCMC) method implemented in the BEAST 1.10.4 tool with BEAGLE library v.3 to accelerate the computational analysis[40,41]. The XML file was built with BEAUTI 1.10.4, using the collection date traits, the non-parametric Bayesian Skyline model, the evolutionary model under GTR+F+G4 and a strict clock with the initial value of substitution rate of 8×10^−4^ substitutions/site/year and continuous-time Markov chain (CTMC) rate reference prior [42,43]. Three runs of 200 million generations were independently performed and the trees combined applying a burnin of 25%, the convergence of analysis was evaluated with TRACER v1.7[44], the maximum clade credibility tree was summarized with TreeAnotator V1.10.4 and the consensus tree was plotted with ggtree[45]. To assess the age root of each clade, a secondary Bayesian analysis was performed using datasets of highly supported clades selected in the previous analysis, each dataset was submitted to the same strategy as the first Bayesian analysis.

### Epidemiological data

SARS-CoV-2 lab confirmed (RT-qPCR) cases and deaths from Brazil and Pernambuco state were recovered from the Brazilian Ministry of Health coronavirus database (https://dados.seplag.pe.gov.br/apps/corona_dados.html). We collected the number of cases and deaths per day and per epidemiological week since the first case reported in the state (March 12th to September 2nd). The data was parsed using dplyr and tidyverse packages and plots were performed using the ggplot2 package of the R statistical language (https://www.r-project.org/).

### Monthly maps of SARS-CoV-2 infection in the state of Pernambuco

We calculated the monthly incidence using the number of cases in the municipality, divided by the estimated population in the year 2020 and multiplied by 100,000 inhabitants. We collected data regarding SARS-CoV-2 cases from the “Covid 19 in Data” platform available on the website of the Planning Department of the State of Pernambuco, on https://dados.seplag.pe.gov.br/apps/corona_data.html. For the construction of the maps, the cartographic base of the municipalities of Pernambuco was collected, in shapefile format, geographic projection system (latitude/longitude), geodetic reference system SIRGAS 2000, available on the IBGE website. Subsequently, to visualize the incidence of SARS-CoV-2 infection in the state of Pernambuco, thematic maps categorized into quartiles were generated.

### Vehicle Mobility Flow Maps in Brazil and Pernambuco

We generated maps of vehicle mobility flow to identify displacement patterns considering the city of origin and destination. The database containing this information was made available through the Origin-Destination Survey of the PNCT (National Traffic Count Plan) by the DNIT (National Department of Infrastructure and Transport) through the link http://servicos.dnit.gov.br/datapnct/SearchOD/Database. We generated a new database to carry out the georeferencing analysis. We considered the following variables: municipality of origin with geographic coordinates, municipality of destination with geographic coordinates, and flow amount. We used the MMQGIS (Hub Lines/Distance) package of the QGIS 3.10 to elaborate the flow maps.

### Highway traffic map in Pernambuco

For spatial modeling of the density flow of vehicles on the highways of the state of Pernambuco, we used a database containing traffic counting information collected through electronic counters positioned at specific points on the highways to cover the most representative sections of the road network in the state of Pernambuco. We recovered this dataset from the National Traffic Count Plan (PNCT) of the National Department of Infrastructure and Transport (DNIT) from http://servicos.dnit.gov.br/dadospnct/ContagemContinua. There are currently 22 electronic counting posts spread across the state of Pernambuco. Based on the information collected, a new database was generated to carry out the georeferencing of the intensity of the flow of vehicles, considering the information referring to geographic coordinates and quantity of vehicles from each monitoring station, being representative for each section of the road network. We also collected from DNIT the digital cartographic database of the state’s highways, as well as the cartographic database of the municipalities of Pernambuco from IBGE website. The database with vehicle traffic information was later added to the shapefile file of highways. To visualize the intensity of the flow of vehicles on the highways of Pernambuco, we used the Ordinary Kriging method. The maps were generated using ArcGIS 10.1 software.

## RESULTS AND DISCUSSION

### Genomic spreading of lineages of SARS-CoV-2 in the Pernambuco State

We obtained 1,389 new genomes with average coverage breadth and depth of 99.65 (stdev 1.57) and 487.27 (stdev 506.99), respectively, encompassing samples collected from 1st June 2020 to 9th August 2021 **(Supplementary Table 1**). The first confirmed SARS-CoV-2 infections in Pernambuco, comprising an elderly couple returning from Italy on 28 February, was reported in the second week of March (12 March 2020) in the 11th epidemiological week, 16 days after the first confirmed case in Brazil (25 February 2020)[46]. Soon after the first case of SARS-CoV-2 infection in the state, we sequenced 101 genomes, from April to mid-May 2020, which all were classified in the B lineage representing multiple introductions of SARS-Cov-2 in the state[31]. The present work is an update of the genomic surveillance comprising the sequencing and analysis of 1,389 new genomes from Pernambuco covering the emergence and spread of at least six epidemiologically important lineages: B1.1, B.1.1.33, B.1.1.28, P.2 (Zeta), (Gamma) and B.1.617.2 - AY.* (Delta) (**Figure 1 and Supplementary Table 1**). The pandemics in Pernambuco state was characterized by two large case incidence and death peaks: one at the first semester of 2020 that stabilized during the second semester and a large peak at the first semester of 2021 (**Figure 2A-B)**. Genomes sequenced in this study covered all months representing about 0.5% of all cases reported in the state.

**Figure 1.**
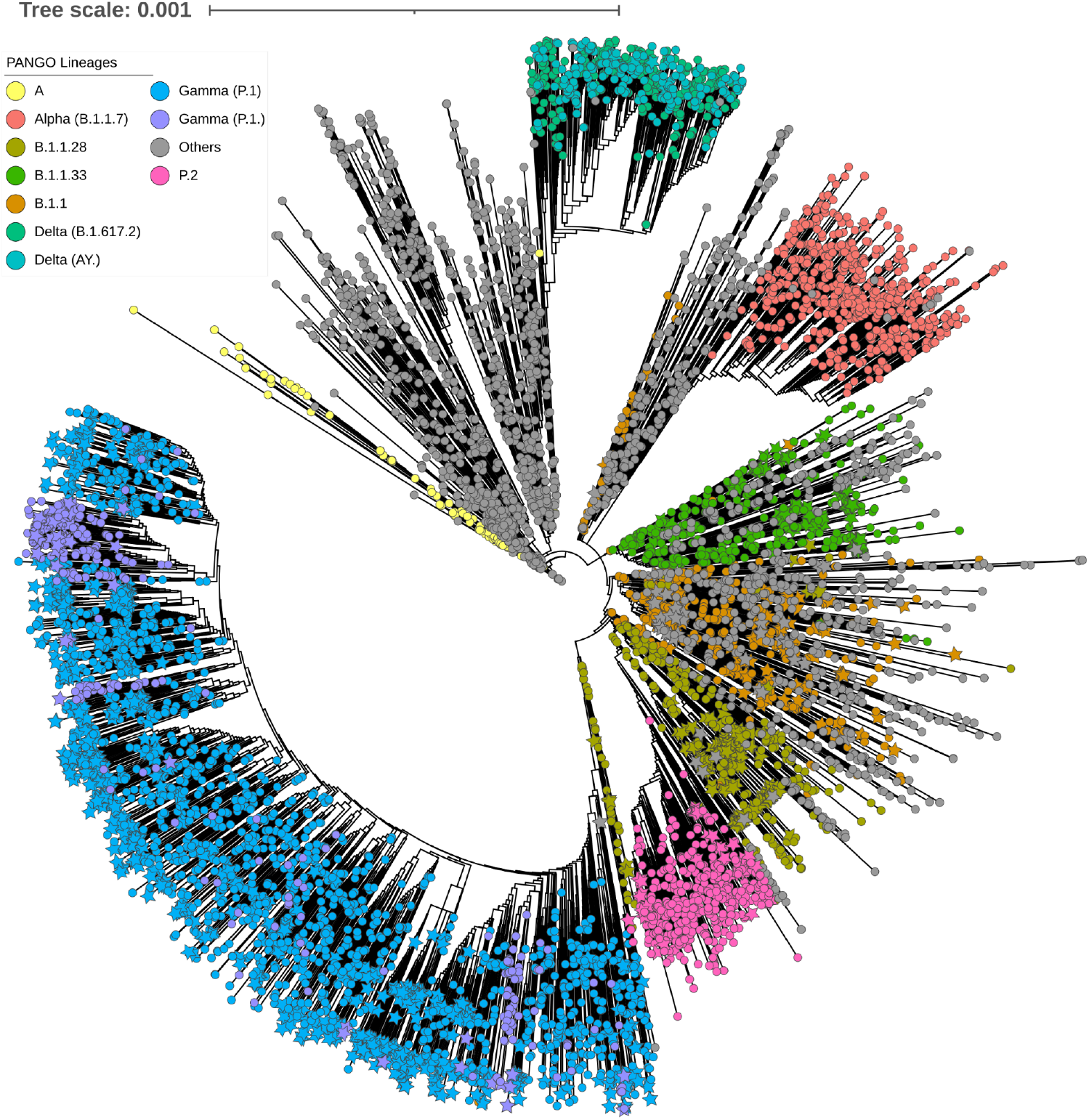
Maximum likelihood phylogeny of SARS-CoV-2 with the global dataset of 7,228 sequences, Pernambuco sequences marked with stars and colors denote lineages groups.

**Figure 2:**
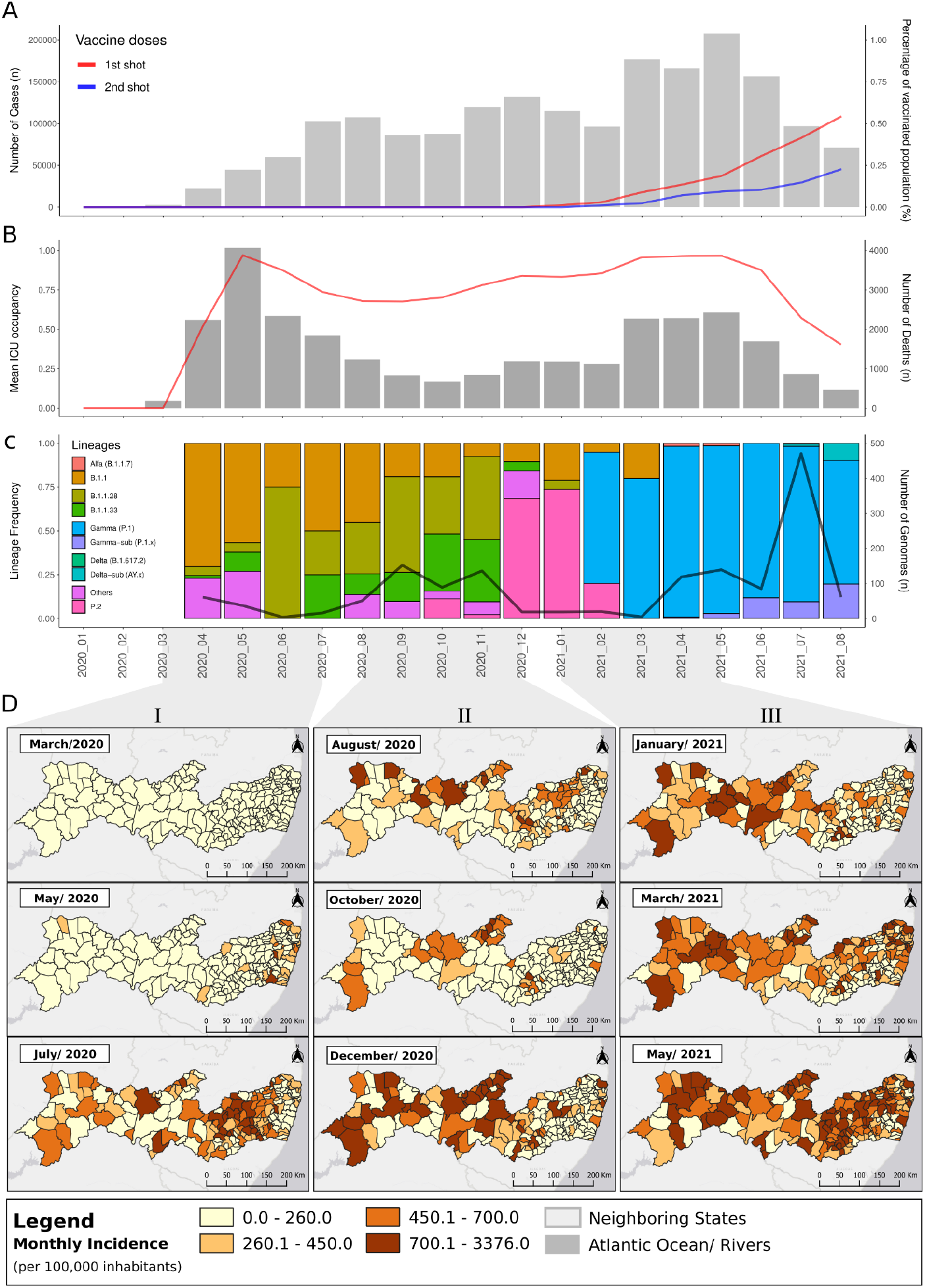
A) Number of SARS-CoV-2 infections and percentage of Pernambuco population vaccinated; B) Patients death (gray bars) and mean ICU occupancy (red line) in Pernambuco by period; C) SARS-CoV-2 lineage frequencies and number of genomes by period; D) Monthly incidence of SARS-CoV-2 infections across Pernambuco State.

Our analyses indicated the most prevalent lineages during the first infection wave at Pernambuco were B.1.1, B.1.1.33, B.1.1.28 and it marked the first wave of SARS-CoV-2 infections between March-May 2020 in the state (**Figure 2C**). During October 2020 the first cases of the lineage P.2 was detected in Pernambuco, at the time P.2 was considered a variant of interest (VOI Zeta) due to specific spike mutations and increasing prevalence in Brazil [32,47]. Nevertheless, the B.1.1, B.1.1.33 and B.1.1.28 lineages remained the most prevalent lineages between October and November 2020. However, between December 2020 and January 2021 the lineage became the most prevalent lineage which was associated with a moderate increase in case numbers and near stabilization of ICU bed occupancy (June-September 2020) (**Figure 2B**). Then, the VOC Gamma (P1) arrived in the state in February and rapidly replaced all previous circulating lineages, becoming the most prevalent lineage from February until August 2021 (**Figure 2C**). These findings are corroborated by the sequencing of the S gene in additional 135 independent Sars-CoV-2 samples (**Supplementary Figure 1**). During this period, case numbers and deaths soared, reaching a highest peak at that point of the pandemic (**Figure 1A and B**). However, during the time frame in which P.2 and P.1 Gamma lineages co-circulated (January-February 2021), the increase in case number was buffered during the transition to Gamma dominance (**Figure 2A and C**).

From January 2021 the vaccination campaign started in the state, but due to a slow rollout, it most likely had a limited impact on cases and deaths during the Gamma wave: as of August 2021, only 25% of the local population were fully vaccinated (**Figure 1A**). Therefore, the stabilization of ICU bed occupancy and deaths (March and May 2021) and its progressive decrease in case numbers after June 2021 is more likely explained by a composite effect of the immunological barrier acquired from natural infection of previous non-VOC lineages, the immunological barrier created by the widespread infection of the population from the Gamma lineage itself (**Figure 1C**) and to the more intense adherence of the population to non-pharmacological intervention (NPI) during high health system pressure [33,48]. Although some of the NPI (Multidisciplinary and multi-sectoral measures, social distancing and border control, obligation of face masks in public environments) were unevenly implemented throughout Pernambuco and for short periods of time, these interventions probably contributed to the reduction of virus transmission in the state. Finally, another VOC, Delta, was introduced to the state in July 2021 and gradually replaced the dominant Gamma lineage without major increase of case and deaths likely due to previous natural and hybrid immunity of the population (**Figure 1C**).

The analysis of the SARS-CoV-2 incidence through time and its association with lineage assignments in different Pernambuco counties revealed three distinct epidemic periods: **I** - March to July 2020 was characterized by a large increase in incidence (incidence peak 398/100.000 inhabitants) mostly driven by the east largest metropolitan region of the State - the Recife Metropolitan Region **(Figure 2D)**. This period of pandemics is characterized by a rapid increase in cases reaching the peak in May 2020 which was dominated by three major cocirculating lineages B.1.1, B.1.1.28 and B.1.1.33. During this same month there is clear evidence of interiorization in which the incidence increased in the west part of the state during June/July 2020 which was followed by extensive transmission of all dominating lineages covering the entire state (**Figure 2D**). The following months (August and September) were characterized by an overall decline in incidence (300∼200 cases per 100.000 habitants) in the entire state (**Figure 2D**). **II** - August to December 2020 was characterized by the emergence of the P.2 lineage in the east region of the state which occurred concomitantly to a new increase in incidence (incidence peak 458 cases by 100,00 inhabitants) which was more pronounced in the west and middle portion of the state (**Figures 2D**). The increase in cases in the west and middle region of the state was likely mainly driven by extensive community transmission of lineages B.1.1.28 and B.1.1.33 and P.2 transmission (**Figure 2D**). However, the low number of genomes obtained during this period may have limited the inferences of lineage prevalence. **III** - January to May 2021 was characterized by the arrival of the P.1 (Gamma) lineage (January) that replaced all previous circulating lineages between February and March and drove the rapid incidence increase all over the state from March 2021 (**Figure 2D**). The peak of infection cases was in May (incidence peak 769 cases per 100.000 inhabitants) followed by a continued decline in cases up to August 2021 (**Figure 2A**). In this same period we also detected the emergence of the Delta lineage in the East part of the state (**Figure 2C**).

### SARS-CoV-2 clades detected in Pernambuco State

We detected fourteen SARS-CoV-2 monophyletic clades containing only Pernambuco samples until the first half of 2021 (**Figure 3**). Our Bayesian analysis showed that the single B.1 clade showed the most ancient last common ancestor (**Figure 3A - upper right**) in the late April to middle May of 2020 (**Table 1**) what is in line with the emergence and predominance of this clade in the first weeks of SARS-CoV-2 transmission in the state from our previous study [31]. Moreover, three different clades of B.1.1.28 co-circulated with two B.1.1.33 clades in the second semester of 2020 - late June to late October (**Table 1**) - showing similar more recent last common ancestors (**Figure 3A - upper right**). Lastly, we detected at least eight clades of P.1 (Gamma) lineage in the state that replaced all previous circulating lineages (**Figure 3A**). This pattern mirrors the P.1 (Gamma) replacements that occurred in other regions of Brazil. For instance, the B.1.1.28 was prevalent in Amazonas state during the second semester of 2020, however shortly after the emergence of the P.1-Gamma lineage it was rapidly replaced by this VOC lineage[49]. Likewise, the same was observed in other Brazilian states [50,51]. After the origin of Gamma lineage on Amazonas state in the late November of 2020 [27], the subsequent spreading to other Brazilian states occurred mainly between December of 2020 and March of 2021 [52]. Current studies estimate the entry of Gamma lineage in the Northeast region varies from the middle-late December of 2020 [52] to early January 2021 [21] which is consistent with the tMRCA of the Gamma lineage in the Pernambuco state (1st December 2020 [95% high posterior density (HPD): 7th November 2020 – 6th January 2021 **Figure 3A**). Regarding the root ages of Gamma clades that seeded large transmission chains in the state, our analysis estimates that they date back between early February 2021 (clade 12, **Table 1**) and mid April 2021 (**Figure 3A, Table 1**).

**Figure 3:**
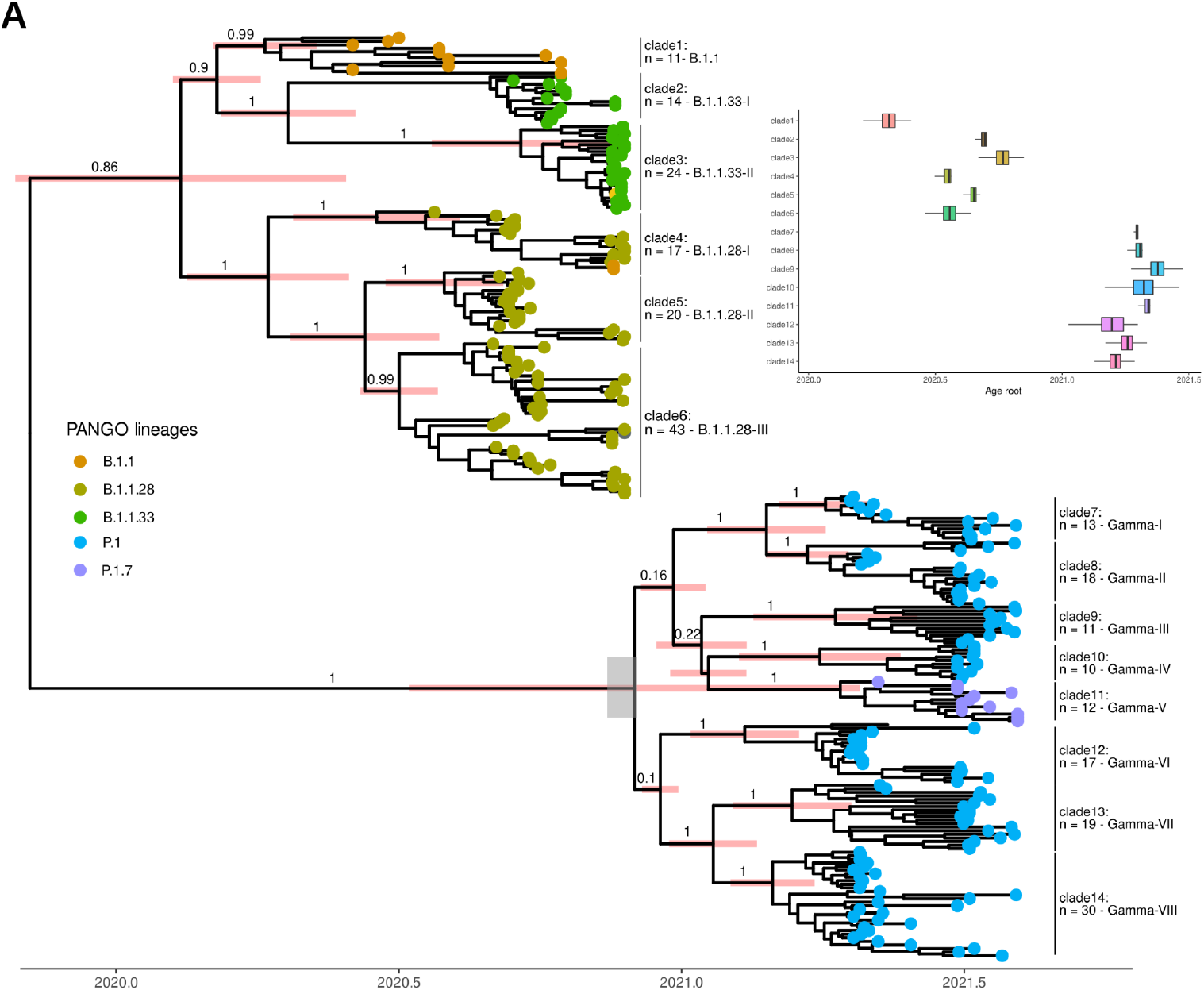
**A**. Temporal analysis of SARS-CoV-2 clades detected in Pernambuco state. Gray rectangle represents the estimated origin of Gamma lineage in Amazonas state (Naveca, et al 2021).

**Table 1:** tMRCA and HPD intervals of each identified clade in figure 3.

| clade | Number of sequences | lineages | tMRCA | 95% HPD min. | 95% HPD max. |
| --- | --- | --- | --- | --- | --- |
| 1 | 11 | B.1.1 | 2020-04-24 | 2020-03-26 | 2020-05-19 |
| 2 | 14 | B.1.1.33 | 2020-09-09 | 2020-08-27 | 2020-09-13 |
| 3 | 24 | B.1.1.33 | 2020-10-05 | 2020-09-08 | 2020-10-28 |
| 4 | 17 | B.1.1.28 | 2020-07-18 | 2020-06-29 | 2020-07-25 |
| 5 | 20 | B.1.1.28 | 2020-08-25 | 2020-08-12 | 2020-09-03 |
| 6 | 43 | B.1.1.28 | 2020-07-21 | 2020-06-23 | 2020-08-13 |
| 7 | 13 | Gama | 2021-04-17 | 2021-04-11 | 2021-04-19 |
| 8 | 18 | Gama | 2021-04-17 | 2021-03-20 | 2021-04-26 |
| 9 | 11 | Gama | 2021-05-16 | 2021-04-16 | 2021-06-12 |
| 10 | 10 | Gama | 2021-04-26 | 2021-03-14 | 2021-06-02 |
| 11 | 12 | Gama | 2021-05-02 | 2021-04-19 | 2021-05-07 |
| 12 | 17 | Gama | 2021-03-12 | 2021-02-01 | 2021-04-19 |
| 13 | 19 | Gama | 2021-04-03 | 2021-03-10 | 2021-04-24 |
| 14 | 30 | Gama | 2021-03-17 | 2021-02-21 | 2021-04-05 |

### Spatio-temporal incidence and inner state dynamics

Overall, we detected a pronounced East-West dispersion pattern from the metropolitan region to the inner part of the state for all lineages, but also a higher concentration of cases in the municipalities that border the states neighboring Pernambuco, indicating a spatial discontinuity with the East-West dispersion pattern. This finding may be related to the presence of main highways that connect these smaller municipalities to other highly-populated regions from the neighboring states. The movement of infected individuals from other states through means of ground transport may have contributed to the further spread of the virus (**Figure 3**). It is important to note that we only used ground traffic data for intra municipalities mobility data in our analysis. Therefore, the results should be evaluated with caution as air transportation may also contribute substantially to respiratory viruses transmission[53,54]. However, intra-state flight mobility in Pernambuco is expected to have a minor impact on human mobility as there are only two large airports in Recife and Petrolina cities, hence, most of the human mobility within the state is expected to occur through highways.

Interestingly, we observed an overlap between the early circulation of new SARS-CoV-2 lineages in discontinued areas of the state and the interstate vehicle mobility flux. This finding can be evidenced on the Western limit of Pernambuco (Chapada do Araripe and Vale do São Francisco regions), which borders the states of Ceará and Bahia and are important highway hubs (**Figure 4A and C**). Moreover, vehicle mobility analysis showed that the metropolitan region of Recife receives an intense flux of vehicles from other Northeastern state capitals and from the cities of Belém in the North and São Paulo, in the Southeast.

**Figure 4:**
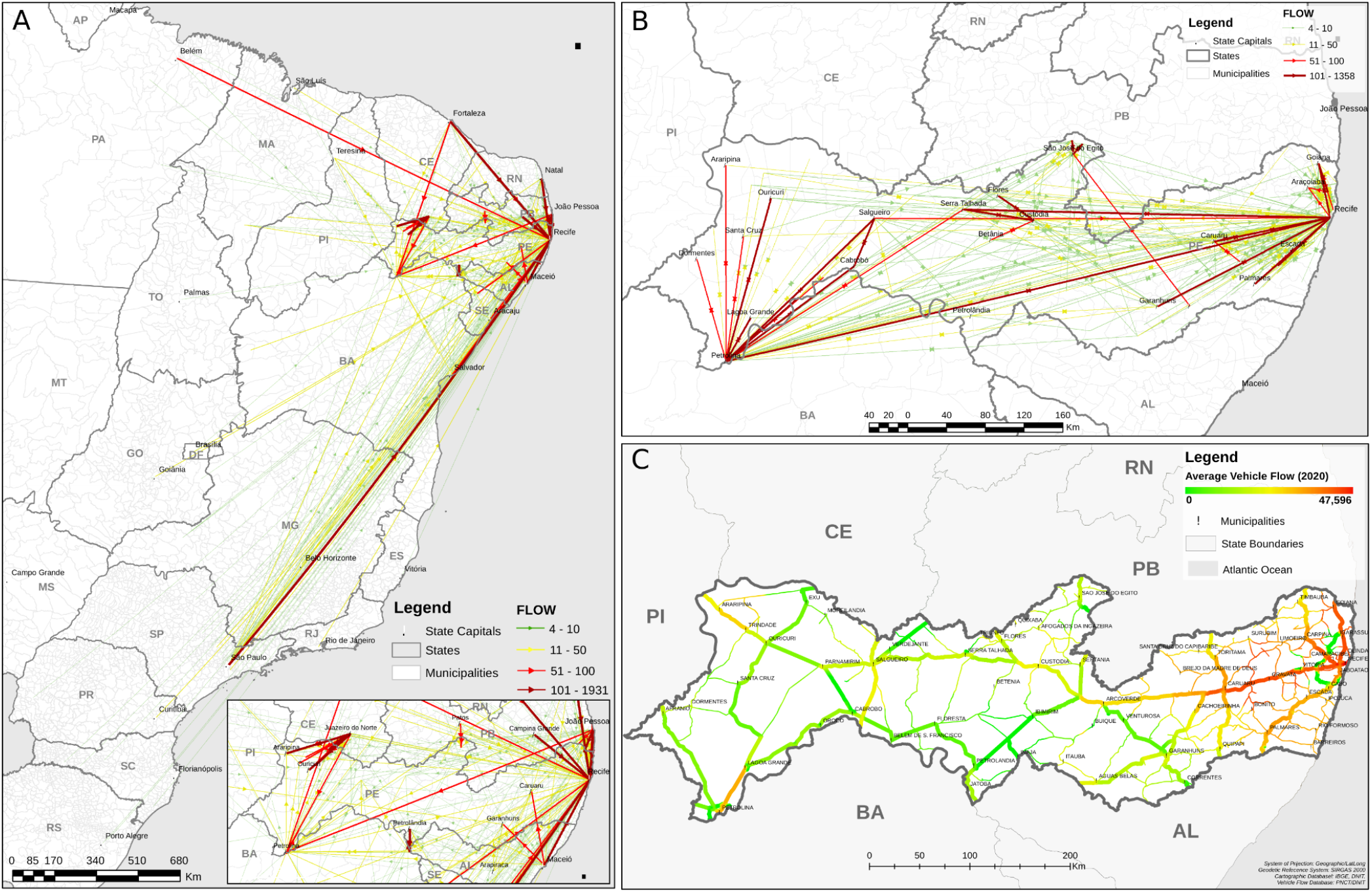
Origin and density of vehicle mobility in the state of Pernambuco. **(A)** shows the origin/destination intensity of vehicles departing from elsewhere in Brazil to the state of Pernambuco. **(B)** shows the destination from vehicles with both origin and destination within the state of Pernambuco. **(C)** shows the modeling of the average 2020 vehicle flux intensity on the roads according to the automated vehicle counters distributed throughout the region.

Corroborating our data, studies conducted by different groups have led to the association of the dispersion of COVID-19 with transportations, such as buses, trains and airplanes[55–57]. A study based on the spatial dispersion of COVID-19 conducted in Bahia, another Brazilian northeastern state, also showed that the disease was distributed across the state through highways and airports[57]. As observed in Recife, the capital of Pernambuco, and where the International Airport is located, the highway network from the Metropolitan Region to the inner cities of the state likely had an important role in spreading the virus towards the country’s municipalities.

### Spike region genotyping analysis with Sanger sequencing

While performing molecular surveillance we also evaluated 135 additional SARS-CoV-2 positive samples by Sanger sequencing to speed up the obtention of actionable results during the emergence of P.1-Gamma lineage in the state. The data obtained in these subset corroborates the rapid P2 and B.1 lineages displacement caused by the Gamma (P.1) variant of concern in Pernambuco between January and April 2021. Of note, in this subset Gamma represented 4.7% of the samples collected in January, 64% in February, 88% in March and 100% in April (**Supplementary Figure 1**).

**Supplementary Figure 1.**
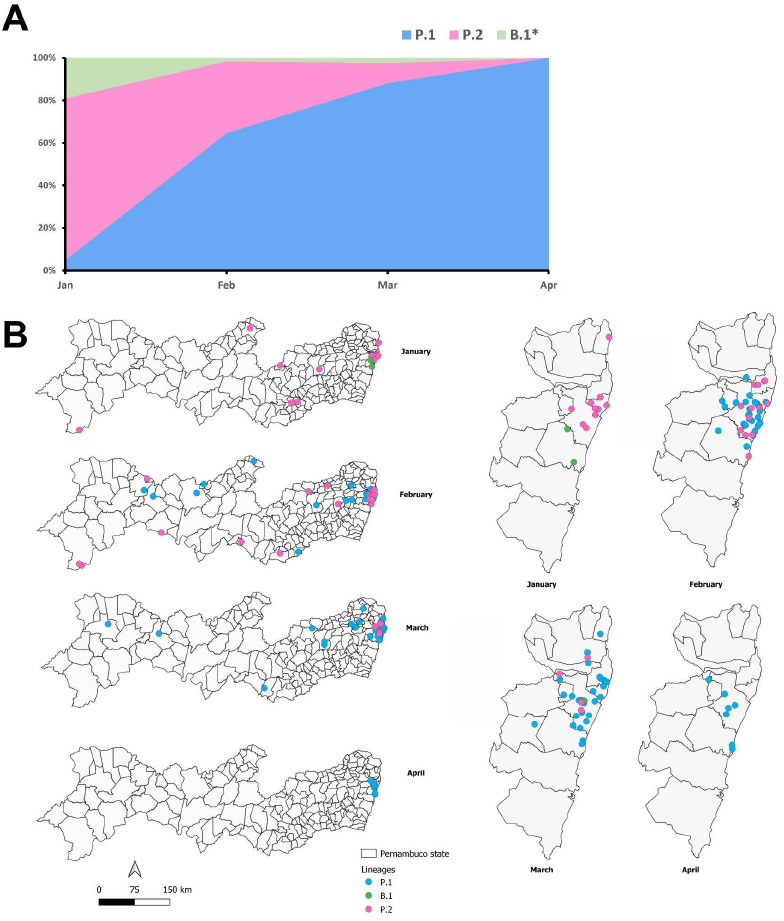
Spatiotemporal distribution of SARS-CoV-2 lineage according to the Sanger sequencing approach. **(A)** The analysis from 135 additional COVID-19 cases through the Sanger sequencing inference protocol shows a fast displacement of P.2 and B.1-derived lineages by the P.1 (Gamma) variant of concern. The maps show the location of each case according to the month of diagnosis. **(B)** The metropolitan region is amplified on the right for a more detailed visualization.

## Conclusion

The SARS-CoV-2 pandemic is one of the most challenging public health emergencies of our time and it remains critical to understand how this virus evolves and spreads at different spatial scales. Here, through viral transmission chain reconstruction from SARS-CoV-2 genomic data and geospatial analysis we characterized three main SARS-CoV-2 transmission waves in the Pernambuco state. Our findings corroborate the multiple lineage circulation dynamics of the first infection wave of the pandemic and the rapid replacement by P.1 (Gamma) lineage, which dominated the second and larger epidemic wave that occurred in the first semester of 2021 and swiped through Brazil. Lastly, transport spatial analysis showed specific inter and intra-state traffic flow patterns that can be leveraged to implement non-pharmacological actions to mitigate further SARS-CoV-2 and other high-impact respiratory viruses transmission in both Pernambuco state and the Northeast region of Brazil.

## Data Availability

All data produced in the present study are available upon reasonable request to the authors

## Acknowledgment

We thank all the researchers that have made SARS-CoV-2 genomes available and the health staff fighting against the COVID-19 pandemics.

